# The exploration of the characteristics of lower extremity deep venous thromboembolism in patients at admission to a tertiary general hospital in a fourth-tier city in China

**DOI:** 10.1101/2024.12.12.24318933

**Authors:** Jianghao Pan, Jianming Sun, Jiaming Song, Guangyin Fu, Yong Lei, Le Wang

## Abstract

With the acceleration of the pace of life and the change of people’s living and working habits, the incidence of venous diseases of lower extremity remains high, and even shows an upward trend year by year, which has a great impact on the quality of life and work efficiency of patients. Well, the occurrence and development of lower extremity deep vein thrombosis (LEDVT) is also related to the economic and humanistic characteristics of a certain place.

**Objectives:** The purpose of this paper is to retrospectively analyze the medical records of DVT patients in a general hospital of a fourth-tier city in China, and find some noteworthy characteristics, so as to provide some objective evidence for the diagnosis of DVT in similar hospitals.

**Methods:** Adult patients with central LEDVT with or without pulmonary embolism (PE) were retrospectively analyzed. The general condition, concomitant diseases, laboratory tests and other clinical characteristics were analyzed.

**Results:** A total of 100 patients’ medical records were analyzed. The age was non-normally distributed (*p* =0.004), which from 31 to 87 (65.3±11.5) years, and 58% of them were 65 years or older. Males (56%) were slightly more than females. Ninety of ninety-four patients’ D-dimer ≥1μgDDU/ml. Fifty-eight were definitely diagnosed with central LEDVT at admission, of which twenty-three were confirmed as PE by CT. Thirty-five patients with leg symptoms, thirty-three patients with chest symptoms, only two patients had both leg and chest symptoms, and thirty-four patients without any obvious symptoms of DVT when they were admitted to the hospital. Only twelve patients were admitted only with LEDVT, and most of the other patients were complicated with other system/organ diseases.

**Conclusions:** LEDVT is more common in elderly patients aged ≥ 65 years. However, the incidence of patients ≥75 years old observed in this study may be limited by the local economic situation and life expectancy. The symptoms can present as many, may be the typical leg symptoms, progression to PE may involve chest symptoms, or it may be neither leg symptoms nor chest symptoms that are masked by the symptoms of other concomitant diseases. LEDVT can be complicated with multiple system/organ diseases. It is necessary to screen for DVT in patients with abnormal D-dimer.

## Introduction

Venous thromboembolism (VT) includes DVT and pulmonary embolism (PE). DVT most often occurs in the leg vein but can also develop in the splanchnic, cerebral, and arm veins. The annual incidence of DVT of the leg is between 48 and 182 per 100,000 in the population[1]. It is the most worrisome of the aetiologies of acute leg swelling and prompt diagnosis and management is essential to minimize the risk of PE and post thrombotic syndrome (PTS). The PTS is a spectrum of signs and symptoms of chronic venous insufficiency, which ranges from mild ankle swelling to debilitating venous claudication or leg ulcers[2]. DVT is a common cause of morbidity and mortality and the third most common cardiovascular disease, after myocardial infarction and stroke. Afflicting worldwide nearly 10 million people of all ethnicities per year, VT is a substantial contributor to the global burden of disease[3]. The annual incidence of acute VT is 1–2 cases per 1000 population[3–5], which rises exponentially with age for both men and women, and is 4-times higher in high-income than low-income countries[6]. The diagnosis of DVT of the leg can be difficult with clinical findings and history being unreliable, though there are so many guidelines on the detection and subsequent management of DVT and suggests the incorporation of a clinical prediction score (Wells score), D-dimer test and venous duplex ultrasound.

Therefore, we hope to further explore the etiology of venous thrombosis of lower extremitys by retrospectively comprehensively analyzing the basic characteristics, disease characteristics, complications and other factors of inpatients with LEDVT in a fourth-tier city, that economic and demographic factors are at the middle level in China, expecting to provide reference information for the future diagnosis and treatment of hospitals at the same level.

## Methods

This study was part of the project “The analysis of the treatment strategy and safety of acute lower extremity deep venous thrombosis” (Medical Science Research Project of Hebei Province, No. 20220459) which was reviewed and approved by the Ethics Committee of Hengshui People’s Hospital, Hebei Province, China (Approval No.: 2021-3-009-1). This part is only a retrospective observation of hospitalized patients, and does not involve disclosure of patients’ personal information and treatment strategies, which are anonymous information. In this part, only the medical records retrieved according to the key words were analyzed. No minors were involved. Personal patient information was anonymized. Therefore, no remedial informed consent was obtained.

### Study Design and Participation

This was a single-center observational study of medical records conducted in a fourth-tier city in China.

### Included and Excluded Criterion

Inclusion criteria: a) age ≥ 18 years, b) patients with an objectively diagnosed, proven episode the central type of LEDVT, including total femoral vein thrombosis and iliac vein thrombosis or a pulmonary embolism (PE) were included, c) hospital stay > 48 h, d) availability of results ultrasonography, e) all patients were diagnosed with DVT for the first time. Exclusion criteria: serious medical problems without tolerating any treatment. No patients were excluded owing to the use of anticoagulation drugs.

### Data Collection and Definitions

Data were collected from January 1 to December 31, 2023. General information, including each patient’s age, gender, the chief complaints, previous medical history, and associated diseases of the enrolled patients were retrospectively retrieved, and the laboratory tests and examination results of the patients in the outpatient clinic, emergency department or after admission were collected.

The results of laboratory tests, which measured the prothrombin time (PT), activated partial thrombin time (APTT), platelet count, fibrinogen (FIB), fibrinogen degradation products (FDP), international normalized ratio (INR), concentrations of plasma D-dimer (Latex immunoturbidimetric was used, LETIA), and blood lipids, were recorded. All laboratory examination results were obtained from Hengshui People’s Hospital, Hebei Province, China, and the results were interpreted according to the reference range set by the laboratory equipment of the hospital. Admission diagnoses were classified according to ICD-11[7].

We used Doppler ultrasonography to diagnose the VT. The diagnostic criteria was the presence of a constant intraluminal filling defect in femoral and iliac veins for central DVT, as well as spiral computer tomography and pulmonary angiography for diagnosis of PE.

Symptoms were categorized into three categories as described by the patients: swelling and/or pain in the lower extremitys was considered leg symptoms; the presence of chest tightness and/or chest pain was considered chest symptoms, and others were unrelated symptoms.

### Statistics Analysis

Data were analyzed between Jan and Mar 2024. Statistical analysis was performed using the SPSS Version 19.0 (SPSS Inc., Chicago, Illinois, USA).

A descriptive statistical analysis was carried out between the general data and current survey results, including the rate, percentage, mean, and standard deviation. The measurement data were analyzed to determine if the data were normally distributed.

## Result

### Patients’ Characteristics

According to the inclusion criteria, a total of 100 patients’ medical records were screened for analysis. The age from 31 to 87 (65.3±11.5) years, and 74% of the patients were 60 years or older, 58% were 65 years or older, and 39% were 70 years or older. The Kolmogorov-Smirnov test showed that the age was non-normally distributed (*p* =0.004), the 65 to 75 age groups accounted for 46% (Fig 1). There were slightly more males (56%) than females (Table 1).

**Fig 1.**
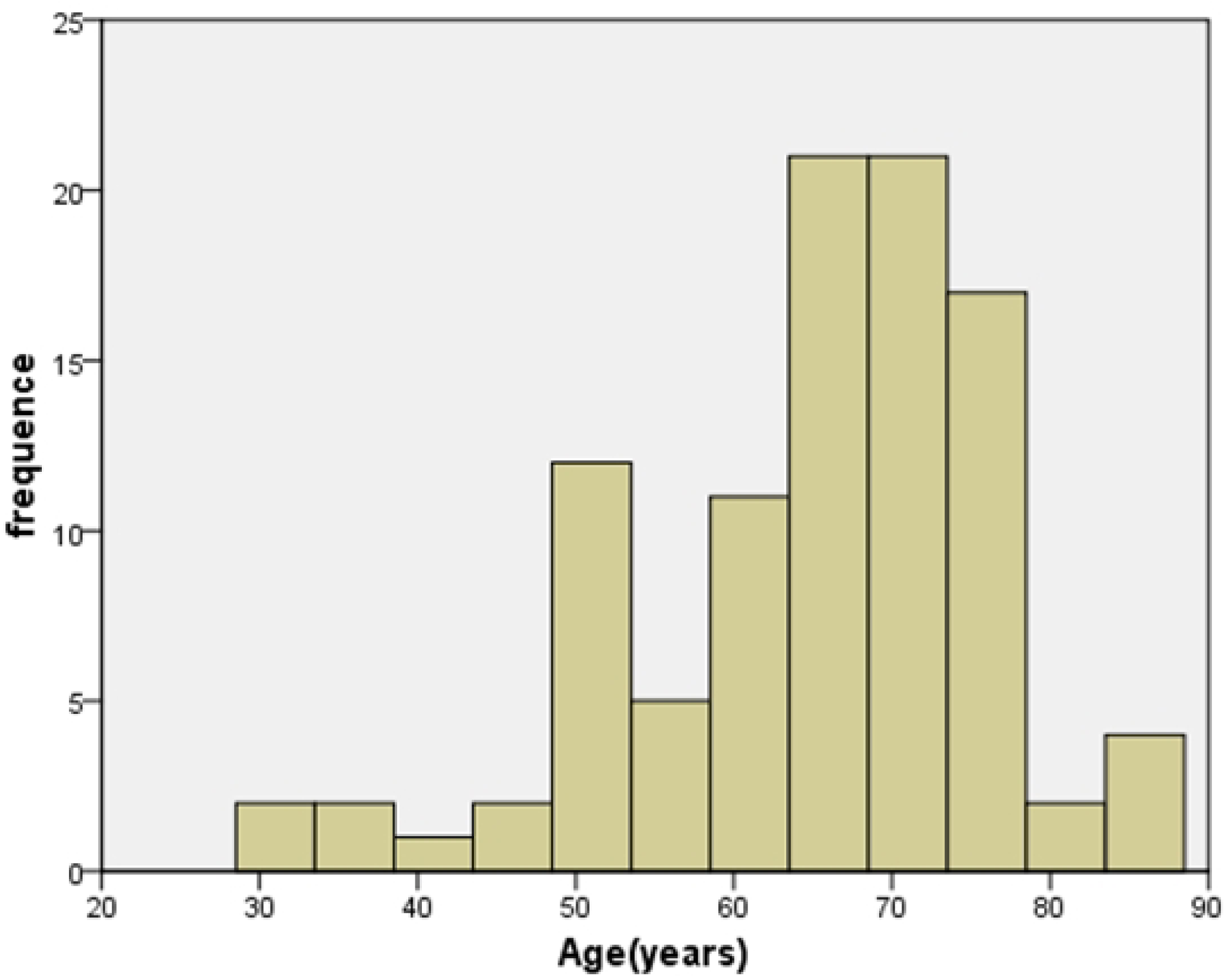
Histogram of all patients according to age distribution.

**Table 1.** Overview of the clinical characteristics of the patients.

| | N | Mean ± SD |
| --- | --- | --- |
| Gender | 100 | - |
| Male (n) | 56 | - |
| Female (n) | 44 | - |
| Age (years) | 100 | 65.30±11.50 |
| $\geq 65$ | 58 | - |
| $< 65$ | 42 | - |
| Temperature ($^{\circ}$C) | 100 | 36.76±2.61 |
| Pluse rate (times/minute) | 100 | 88.05±19.18 |
| $\geq 100$ | 24 | - |
| Respiratory rate (times/minute) | 100 | 19.45±2.68 |
| SBP (mmHg) | 100 | 130.19±22.55 |
| $\geq 120$ | 76 | - |
| DBP (mmHg) | 100 | 79.71±15.07 |
| $\geq 90$ | 24 | - |
| Blood platelet ($\times 10^9$/L) | 99 | 208.83±93.67 |
| ≥300 | 13 | - |
| Blood glucose (mmol/L) | 94 | 8.49±5.00 |
| ≥6.1 | 56 | - |
| Triglyceride (TG, mmol/L) | 72 | 1.61±2.02 |
| ≥1.7 | 17 | - |
| Total cholesterol (TC, mmol/L) | 72 | 4.43±1.30 |
| ≥5.7 | 9 | - |
| Serum creatinine (umol/L) | 100 | 87.12±85.58 |
| ≥81 | 36 | - |
| Blood uric acid (umol/L) | 100 | 336.66±138.85 |
| ≥363 | 34 | - |
| D-dimer (μgDDU/ml) | 94 | 9.72±14.19 |
| ≥1 | 90 | - |
| Prothrombin time (PT, second) | 98 | 11.88±1.52 |
| 9~13 | 89 | - |
| ≥13 | 9 | - |
| Activated partial thromboplastin time (APTT, second) | 98 | 27.32±7.93 |
| ≤25 | 29 | - |
| 25~31.3 | 59 | - |
| ≥31.3 | 10 | - |
| Fibrinogen (FIB, g/L) | 97 | 3.60±1.63 |
| ≤2 | 13 | - |
| 2~4 | 52 | - |
| ≥4 | 32 | - |
| Fibrinogen degradation products (FDP, μg/ml) | 78 | 33.93±51.99 |
| ≥5 | 66 |  |
| International normalized ratio (INR) | 75 | 1.1584±1.09 |
| ≤0.8 | 1 | - |
| 0.8~1.2 | 67 | - |
| ≥1.2 | 7 | - |
| Pulmonary embolism ( n ) | 100 | - |
| Confirmed PE by CT ( n ) | 23 | - |
| uncheck ( n ) | 77 | - |
| Complaint | 102 | - |
| Unrelated symptoms ( n ) | 34 | - |
| leg symptoms ( n ) | 35 | - |
| Chest symptoms ( n ) | 33 | - |

Twenty-four patients had a resting heart rate of more than 100 beats per minute. Seventy-six patients had SBP ≥120mmHg and twenty-four had DBP ≥ 90mmHg. There were fifty-six patients with fasting blood glucose higher than 6.1mmol/L, thirty-six patients with serum creatinine and thirty-four patients with uric acid higher than the upper limit of our hospital detection standard. Triglyceride and total cholesterol were higher than the upper limit of our hospital detection standard in seventeen cases and nine patients, respectively. Ninety of ninety-four patients’ D-dimer had abnormal results (≥1μgDDU/ml). Most of the other coagulation-related indexes (including platelet count) were in the normal range (Table 1).

Although all the enrolled patients had central LEDVT, at the time of admission, forty-two had been admitted because of other medical conditions or discomfort, and only fifty-eight were diagnosed with LEDVT, of which twenty-three were confirmed as PE by CT. Thirty-four patients did not have any obvious symptoms of DVT at admission. Thirty-five patients were admitted to the hospital with the chief complaint of leg symptoms. Thirty-three patients were admitted with chest symptoms as the chief complaint. Only 2 patients had both leg and chest symptoms (Table 1 and Table 2).

**Table 2.** Distribution of symptoms and age ≥ 65 years among system diseases at admission.

| Classification of Diseases | Number | Age $\geq 65$<br>years<br>(n) | unrelated<br>symptoms<br>(n) | leg<br>symptoms<br>(n) | chest<br>symptoms<br>(n) |
| --- | --- | --- | --- | --- | --- |
| LEDVT at admission | 58 | 35 | 8 | 31 | 19 |
| Confirmed PE by CT | 23 | - | 1 | 1 | 21 |
| Hypertensive diseases | 51 | 29 | 18 | 12 | 21 |
| Injury, poisoning or certain other<br>consequences of external causes | 33 | 22 | 15 | 8 | 10 |
| Other diseases of the circulatory system | 31 | 21 | 7 | 8 | 16 |
| Endocrine, nutritional or metabolic diseases (Diabetes mellitus) | 31 | 16 | 10 | 9 | 12 |
| Diseases of the respiratory system | 28 | 14 | 15 | 7 | 6 |
| Cerebrovascular diseases | 26 | 17 | 11 | 5 | 10 |
| Neoplasms | 18 | 12 | 9 | 6 | 3 |
| Diseases of the digestive system | 18 | 12 | 12 | 3 | 3 |
| Symptoms, signs or clinical findings, not elsewhere classified | 15 | 9 | 6 | 4 | 5 |
| Diseases of the nervous system | 12 | 7 | 9 | 2 | 1 |
| Diseases of the musculoskeletal system or connective tissue | 10 | 7 | 2 | 3 | 5 |
| Diseases of the genitourinary system | 5 | 2 | 3 | 1 | 1 |
| Certain infectious or parasitic diseases | 3 | 2 | 1 | 1 | 1 |
| Diseases of the blood or blood-forming organs | 1 | 0 | 1 | 0 | 0 |
| Mental, behavioural or neurodevelopmental disorders | 1 | 1 | 1 | 0 | 0 |

Two patients were diagnosed with coronary artery disease at admission because of chest tightness and pain. Further examination after admission confirmed DVT of the lower extremities and PE. Seven of the twenty-three patients with PE diagnosed by CT had a pulse rate ≥100 beats/min at admission.

Among the 100 patients, twelve patients had no other systemic diseases, fifty-one patients had hypertensive disease, thirty-three patients had injury, poisoning or certain other consequences of external causes, thirty-one had diabetes mellitus, thirty-one had other diseases of the circulatory system (excluding VT and hypertension), twenty-eight had disease of respiratory system, twenty-six had cerebrovascular diseases, eighteen had neoplasms (including cysts and polyps), eighteen had diseases of the digestive system, twelve had diseases of the nervous system (excluding cerebrovascular diseases), ten had diseases of the musculoskeletal system or connective tissue, and five had diseases of the genitourinary system (Table 2).

Among the fifty-eight patients who were diagnosed with LEDVT at admission, thirty-five patients were ≥ 65 years old, thirty-one patients had leg symptoms, nineteen patients had symptoms of chest symptoms, and eight patients had no DVT related symptoms. Twenty-one patients were diagnosed with PE by CT examination because of chest symptoms. Among the patients who were clearly diagnosed with LEDVT at admission, the elderly patients aged ≥ 65 years accounted for the majority of the concomitant diseases in each system. Patients with hypertensive disease, other diseases of the circulatory system and diabetes mellitus had relatively more chest symptoms, which were twenty-one, sixteen and twelve cases, respectively. Most of the patients with other system diseases combined with LEDVT had no related symptoms at admission (Table 2).

It is worth mentioning that one female patient died on the sixth day of admission. The patient was hospitalized with cerebral thrombosis due to poor movement of the left extremity for 2 days. After admission, ultrasound examination showed heterogeneous echo and filling defects in the left common femoral vein, femoral vein, deep femoral vein, popliteal vein, posterior tibial vein, and peroneal vein. Before death, she suddenly developed vertigo, palpitations, and chest tightness while lying down in a sitting position. At the same time, the SPO_2_ decreased to 84%, and the blood pressure was 91/52 mmHg. After 2.5 hours of rescue, there was no relief of symptoms. Blood pressure and SPO_2_ continued to decrease until there was no spontaneous rhythm and respiration, and blood pressure could not be measured. The patient was declared clinically dead.

## Discussion

Although there have been many studies on the causes, influencing factors and diagnostic methods of LEDVT, few papers have summarized the characteristics of patients with LEDVT at admission involving the same level of cities and hospitals.

In this study, most of the patients diagnosed with LEDVT at admission or during hospitalization were elderly. This is consistent with previous studies. As we know the risk of VT increases with age[8]; 60% of all events occur after the age of 60 years. The incidence of VT is less than 5 per 100 000 individuals per year in people younger than 15 years but is approximately 500 patients per 100 000 individuals per year at the age of 80 years[9]. The main age group of patients in this study was between 65 to 75 years old. Although there appears to be a trend toward an increase in LEDVT with age up to 75 years, there is not a continuous increase in patients after age 75 years. Taking into account the local health economic situation, the average life expectancy of residents is related.

Similar to many previous studies, although the patients in this study were all patients with LEDVT, only one third of the patients had typical LEDVT related leg symptoms, and one third of the patients had chest symptoms at admission. As we know the typical clinical signs and symptoms of LEDVT include leg pain (80–90% of patients), swelling (80%), redness (25%), localised tenderness on palpation (75–85%), and prominent collateral superficial veins (30%)[10, 11]. Of the fifty-eight patients with LEDVT diagnosed at admission, thirty-one had definite leg symptoms. Chest symptoms maybe the most obvious and easily recognized symptoms associated with PE. In this study, twenty-one of the twenty-three patients who were diagnosed with PE by CT scan were diagnosed with PE because they had obvious chest symptoms. Take a look at past reports that most patients with PE present with breathlessness (80% of patients), pleuritic chest pain (60–70%), haemoptysis (5–13%), tachycardia (65–70%), or hypoxemia (70%), but can also present with severe haemodynamic compromise (10– 20%), including sudden death, shock, hypotension, syncope, or confusion[12]. The death case involved in this study were not confirmed to be PE by CT, the disease progression and hemodynamic changes during the rescue process were consistent with the contents of these literatures. So the cause of death was highly suspected to be PE. As seen in the study, patients with coexisting hypertension, coronary heart disease, and diabetes are more likely to have chest symptoms. Therefore, when these patients with chest symptoms, it is also crucial to distinguish them from PE. We should note that between 30% to 60% of patients with symptomatic proximal DVT have silent PE[13]. About 40% of patients with symptomatic PE have proximal DVT and 25% have only distal DVT[14, 15]. However, only half of patients with PE and imaging confirmed LEDVT have leg symptoms[16, 17]. Two patients in this study had no leg symptoms and were admitted with chest tightness and/or chest pain as the first symptom. Coronary heart disease was diagnosed at the time of admission, and PE was confirmed only by further examination. Clinically, such patients are indeed easily confused with patients with coronary heart disease, and the differential diagnosis needs to be made quickly in a short time. The harm of PE is self-evident, so it is necessary to perform PE screening in patients with LEDVT if they are eligible.

The majority of patients in the study had coexisting diseases. There were eight patients in this study who were diagnosed with LEDVT at admission had neither leg nor chest symptoms. These patients were found to have LEDVT during the course of the examination for other diseases. And there were thirty-two patients been found to have LEDVT or PE during the diagnosis of other diseases inhospital. There was 15.3% prevalence of DVT in bedridden hospitalized orthopaedic patients[18]. Considering that some patients who require bedridden hospitalization would be potential LEDVT patients. Moreover, the majority of patients involved in this study with D-dimer testing were positive. Screening of these patients is indeed necessary.

Twenty-four of the patients in this study had tachycardia (pulse rate > 100 beats per minute) at admission, and since most of the study patients did not undergo CT. Thus, only 7 patients with confirmed PE had a pulse rate ≥100 beats per minute. the relationship between pulse rate with PE needs to be further explored.

More than half of the patients had blood glucose ≥ 6.1mmol/L at admission. In addition to patients with diabetes, some patients were admitted to the emergency room and were in the non-fasting state when the index was detected.

Similar to other observational studies, this study further confirmed the results of some previous studies, but it inevitably has some limitations. The study only looked at the characteristics of patients diagnosed with central LEDVT at the time of admission, it could not determine the relationship between these characteristics and LEDVT. Due to the single-center study, small sample size and trivial data, it is not easy to systematize and the degree of universality is not high. This study was conducted under natural diagnosis and treatment conditions and is susceptible to interference by various incidental factors, which may affect the accuracy and reliability of the study. This study mainly observed the characteristics of patients with LEDVT at the time of admission, and no further treatment of DVT has been recorded, which makes many attributes and characteristics of the disease not revealed, thus exposing certain limitations.

## Conclusion

LEDVT is more common in elderly patients aged ≥ 65 years. However, the incidence of patients ≥75 years old observed in this study may be limited by the local economic situation and life expectancy. The symptoms can present as many, may be the typical leg symptoms, progression to PE may involve chest symptoms, or it may be neither leg symptoms nor chest symptoms that are masked by the symptoms of other concomitant diseases. LEDVT can be complicated with multiple system/organ diseases. It is necessary to screen for LEDVT in patients with abnormal D-dimer.

## Author Contributions

**Conceptualization:** Jianghao Pan, Jianming Sun, Le Wang.

**Data curation:** Le wang, Jianming Sun, Guangyin Fu, Yong Lei.

**Formal analysis:** Jianghao Pan, Le Wang.

**Project administration:** Jianghao Pan, Le Wang.

**Software:** Jiamin Song.

**Investigation:** Jianghao Pan, Jianming Sun.

**Writing – original draft:** Le Wang, Jianghao Pan.

**Writing – review & editing:** Le Wang, Jianghao Pan, Guangyin Fu, Yong Lei.

## Data Availability

All relevant data are within the manuscript and its Supporting Information files.

